# Epidemic analysis of COVID-19 in China by dynamical modeling

**DOI:** 10.1101/2020.02.16.20023465

**Authors:** Liangrong Peng, Wuyue Yang, Dongyan Zhang, Changjing Zhuge, Liu Hong

## Abstract

The outbreak of novel coronavirus-caused pneumonia (COVID-19) in Wuhan has attracted worldwide attention. Here, we propose a generalized SEIR model to analyze this epidemic. Based on the public data of National Health Commission of China from Jan. 20th to Feb. 9th, 2020, we reliably estimate key epidemic parameters and make predictions on the inflection point and possible ending time for 5 different regions. According to optimistic estimation, the epidemics in Beijing and Shanghai will end soon within two weeks, while for most part of China, including the majority of cities in Hubei province, the success of anti-epidemic will be no later than the middle of March. The situation in Wuhan is still very severe, at least based on public data until Feb. 15th. We expect it will end up at the beginning of April. Moreover, by inverse inference, we find the outbreak of COVID-19 in Mainland, Hubei province and Wuhan all can be dated back to the end of December 2019, and the doubling time is around two days at the early stage.

## 1. INTRODUCTION

A novel coronavirus, formerly called 2019-nCoV, or SARS-CoV-2 by ICTV (severe acute respiratory syndrome coronavirus 2, by the International Committee on Taxonomy of Viruses) caused an outbreak of atypical pneumonia, now officially called COVID-19 by WHO (coronavirus disease 2019, by World Health Organization) first in Wuhan, Hubei province in Dec., 2019 and then rapidly spread out in the whole China^1^. As of 24:00 Feb. 13th, 2020 (Beijing Time), there are over 60, 000 reported cases (including more than 1, 000 death report) in China, among which, over 80% are from Hubei province and over 50% from Wuhan city, the capital of Hubei province^2,3^.

The central government of China as well as all local governments, including Hubei, has tightened preventive measures to curb the spreading of COVID-19 since Jan. 2020. Many cities in Hubei province have been locked down and many measures, such as tracing close contacts, quarantining infected cases, promoting social consensus on self-protection like wearing face mask in public area, *etc*. However, until the finishing of this manuscript, the epidemic is still ongoing and the daily confirmed cases maintain at a high level.

During this anti-epidemic battle, besides medical and biological research, theoretical studies based on either statistics or mathematical modeling may also play a non-negligible role in understanding the epidemic characteristics of the outbreak, in forecasting the inflection point and ending time, and in deciding the measures to curb the spreading.

For this purpose, in the early stage many efforts have been devoted to estimate key epidemic parameters, such as the basic reproduction number, doubling time and serial interval, in which the statistics models are mainly used^4–9^. Due to the limitation of detection methods and restricted diagnostic criteria, asymptomatic or mild patients are possibly excluded from the confirmed cases. To this end, some methods have been proposed to estimate untraced contacts^10^, undetected international cases^11^, or the actual infected cases in Wuhan and Hubei province based on statistics models^12^, or the epidemic outside Hubei province and overseas^6,13–15^. With the improvement of clinic treatment of patients as well as more strict methods stepped up for containing the spread, many researchers investigate the effect of such changes by statistical reasoning^16,17^ and stochastic simulation^18,19^.

Compared with statistics methods^20,21^, mathematical modeling based on dynamical equations^15,22–24^ receive relatively less attention, though they can provide more detailed mechanism for the epidemic dynamics. Among them, the classical *susceptible exposed infectious recovered model* (SEIR) is the most widely adopted one for characterizing the epidemic of COVID-19 outbreak in both China and other countries^25^. Based on SEIR model, one can also assess the effectiveness of various measures since the outbreak^23,24,26–28^, which seems to be a difficult task for general statistics methods. SEIR model was also utilized to compare the effects of lock-down of Hubei province on the transmission dynamics in Wuhan and Beijing^29^. As the dynamical model can reach interpretable conclusions on the outbreak, a cascade of SEIR models are developed to simulate the processes of transmission from infection source, hosts, reservoir to human^30^. There are also notable generalizations of SEIR model for evaluation of the transmission risk and prediction of patient number, in which model, each group is divided into two subpopulations, the quarantined and unquarantined^23,24^. The extension of classical SEIR model with delays^31,32^ is another routine to simulate the incubation period and the period before recovery. However, due to the lack of official data and the change of diagnostic caliber in the early stage of the outbreak, most early published models were either too complicated to avoid the overfitting problem, or the parameters were estimated based on limited and less accurate data, resulting in questionable predictions.

In this work, we carefully collect the epidemic data from the authoritative sources: the National, provincial and municipal Health Commissions of China (abbreviated as NHC, see *e*.*g*. http://www.nhc.gov.cn/) until the article is completed (Feb. 16th, 2020). Then we follow the routine of dynamical modeling and focus on the epidemic of COVID-19 in five most interested regions in China, *i*.*e*. the Mainland excluding Hubei province (denoted as Mainland*), Hubei province excluding Wuhan city (Hubei*), Wuhan, Beijing and Shanghai. Such a design aims to minimize the influence of Hubei province and Wuhan city on the data set due to their extremely large infected populations compared to other regions. Without further specific mention, these conventions will be adopted thorough the whole paper.

By generalizing the classical SEIR model, *e*.*g*. introducing a new quarantined state and considering the effect of preventive measures, key epidemic parameters for COVID-19, like the latent time, quarantine time and basic reproduction number are determined in a relatively reliable way. The widely interested inflection point, ending time and total infected cases in hot cities and regions are predicted and validated through both direct and indirect evidences. Furthermore, by inverse inference, the starting date of this outbreak are estimated. The analysis of other hot spots in China, as well as overseas countries are still in progress.

## 2. MODEL AND METHODS

### A. Generalized SEIR model

To characterize the epidemic of COVID-19 which outbroke in Wuhan at the end of 2019, we generalize the classical SEIR model^23–29^ by introducing seven different states, *i*.*e*. {*S*(*t*), *P* (*t*), *E*(*t*), *I*(*t*), *Q*(*t*), *R*(*t*), *D*(*t*)} denoting at time *t* the respective number of the *susceptible cases, insusceptible cases, exposed cases* (infected but not yet be infectious, in a latent period), *infectious cases* (with infectious capacity and not yet be quarantined), *quarantined cases* (confirmed and infected), *recovered cases* and *closed cases* (or death). The adding of a new quarantined sate is driven by data, which together with the recovery state takes replace of the original *R* state in the classical SEIR model. Their relations are given in Fig. 1 and characterized by a group of ordinary differential equations (or difference equations if we consider discrete time, see SI). Constant *N* = *S* + *P* + *E* + *I* + *Q* + *R* + *D* is the total population in a certain region. The coefficients {*α, β, γ*^*−*1^, *δ*^*−*1^, *λ*(*t*), *κ*(*t*)} represent the protection rate, infection rate, average latent time, average quarantine time, cure rate, and mortality rate, separately. Especially, to take the improvement of public health into account, such as promoting wearing face masks, more effective contact tracing and more strict locking-down of communities, we assume that the susceptible population is stably decreasing and thus introduce a positive protection rate *α* into the model. In this case, the basic reproduction number becomes *BRN* = *β * δ*^*−*1^ *\** (1 *− α*)^*T*^, *T* is the number of days.

**FIG. 1.**
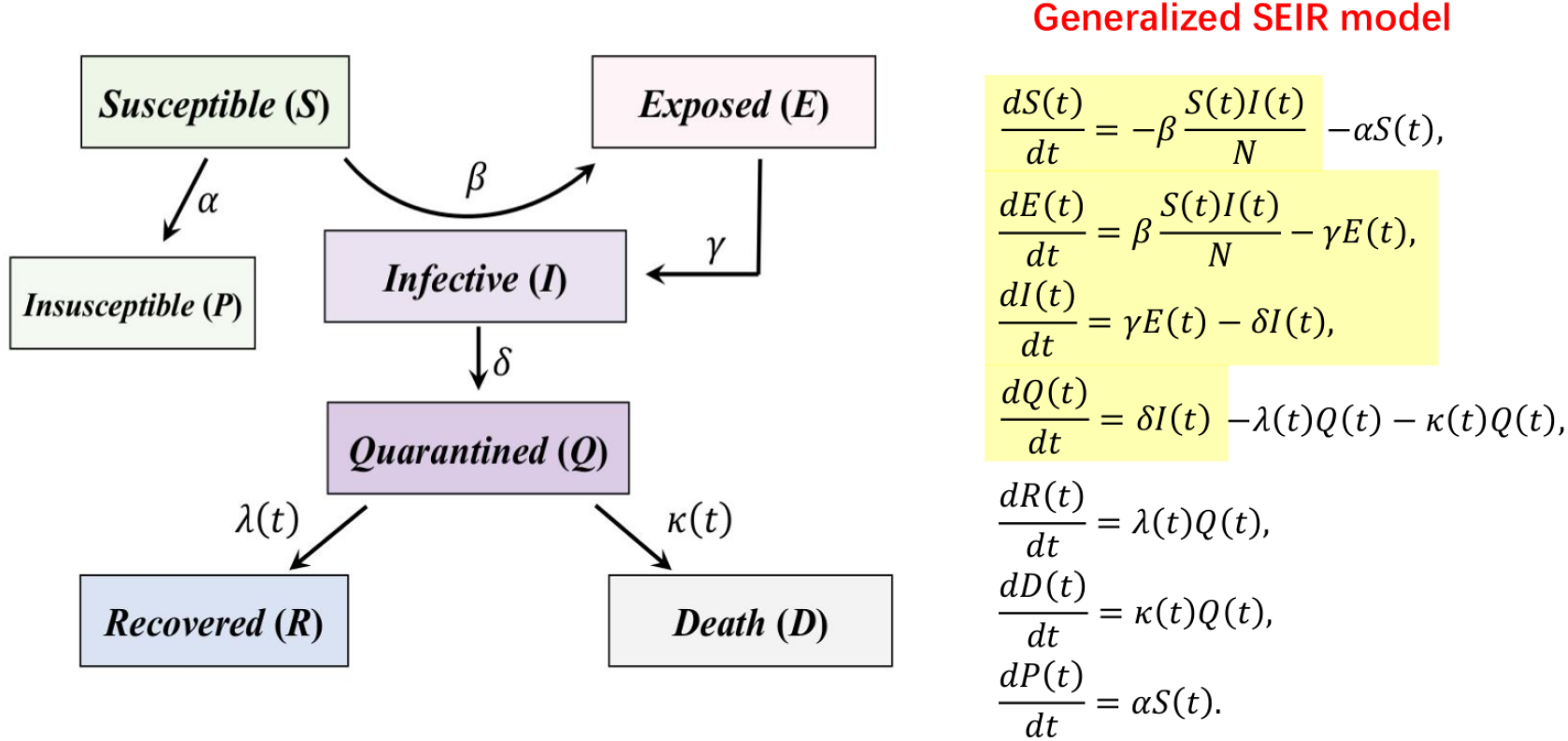
The epidemic model for COVID-19. The highlighted part shows the classical SEIR model.

It is noted that here we assume the cure rate *λ* and the mortality rate *κ* are both time dependent. As confirmed in Fig. 2a-d, the cure rate *λ*(*t*) is gradually increasing with the time, while the mortality rate *κ*(*t*) quickly decreases to less than 1% and becomes stabilized after Jan. 30th. This phenomenon is likely raised by the assistance of other emergency medical teams, the application of new drugs, *etc*. Furthermore, the average contact number of an infectious person is calculated in Fig. 2e-f and could provide some clue on the infection rate. It is clearly seen that the average contact number is basically stable over time, but shows a remarkable difference among various regions, which could be attributed to different quarantine policies and implements inside and outside Hubei (or Wuhan), since a less severe region is more likely to inquiry the close contacts of a confirmed case. A similar regional difference is observed for the severe condition rate too. In Fig. 2g-h, Hubei and Wuhan overall show a much higher severe condition rate than Shanghai. Although it is generally expected that the patients need a period of time to become infectious, to be quarantined, or to be recovered from illness, but we do not find a strong evidence for the necessity of including time delay (see SI for more details). As a result, the time-delayed equations are not considered in the current work for simplicity.

**FIG. 2.**
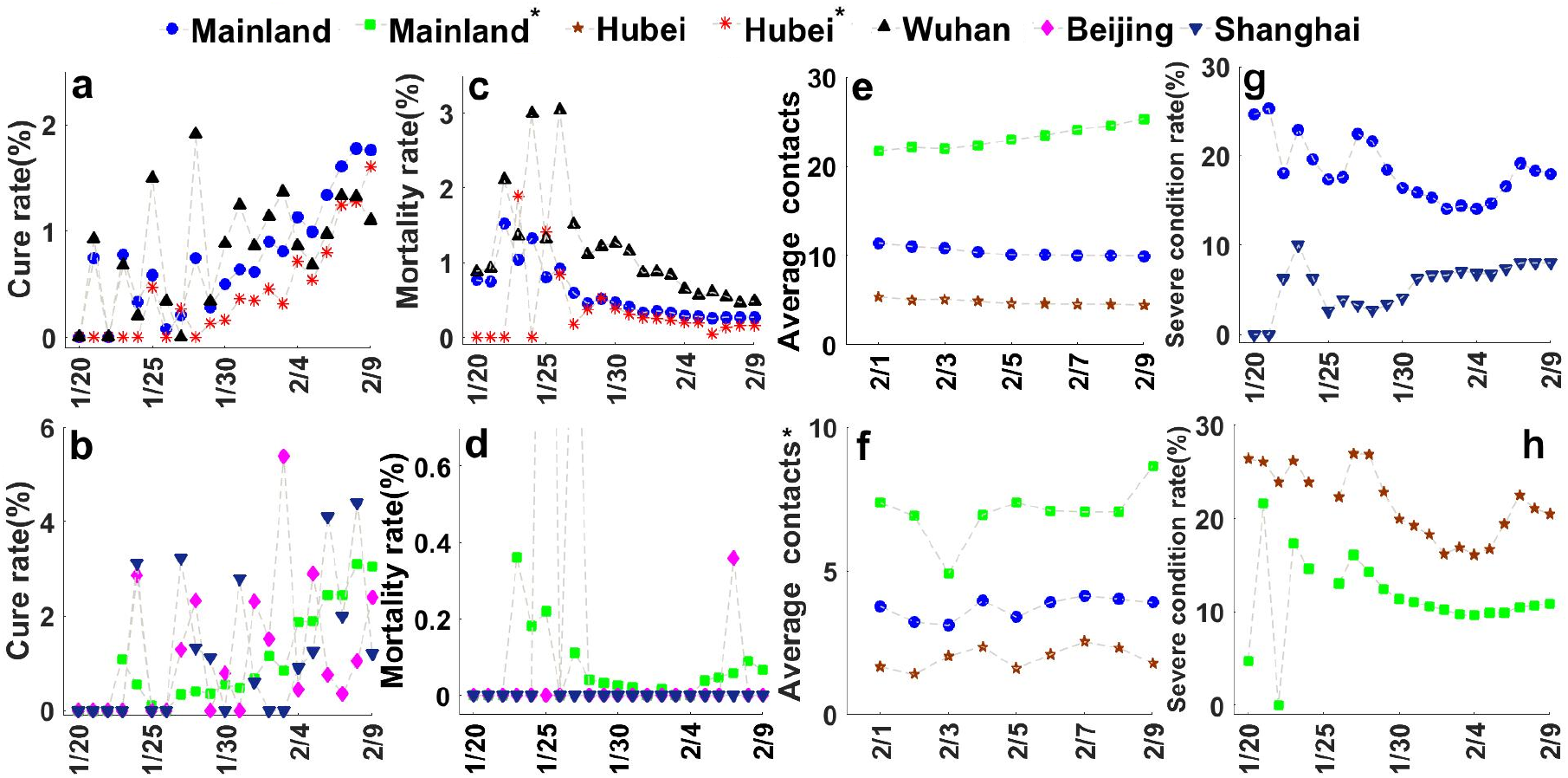
(Color online) (a)-(b) The cure rate *λ*, (c)-(d) mortality rate *κ*, (e)-(f) average close contacts, and (g)-(h) severe condition rate (see SI for their definitions) are calculated based on the public data from NHC of China from Jan. 20th to Feb. 9th for Mainland, Mainland*, Hubei, Hubei*, Wuhan, Beijing and Shanghai separately.

### B. Parameter estimation

According to the daily official reports of NHC of China, the cumulative numbers of quarantined cases, recovered cases and closed cases are available in public. However, since the latter two are directly related to the first one through the time dependent recovery rate and mortality rate, the numbers of quarantined cases *Q*(*t*) plays a key role in our modeling. A similar argument applies to the number of insusceptible cases too. Furthermore, as the accurate numbers of exposed cases and infectious cases are very hard to determine, they will be treated as hidden variables during the study.

Leaving alone the time dependent parameters *λ*(*t*) and *κ*(*t*), there are four unknown coefficients {*α, β, γ*^*−*1^, *δ*^*−*1^} and two initial conditions {*E*_0_, *I*_0_} about the hidden variables (other initial conditions are known from the data) have to be extracted from the time series data {*Q*(*t*)}. Such an optimization problem could be solved automatically by using the simulating annealing algorithm (see SI for details). A major difficulty is how to overcome the overfitting problem.

To this end, we firstly prefix the latent time *γ*^*−*1^, which is generally estimated within several days^5,33,34^. And then for each fixed *γ*^*−*1^, we explore its influence on other parameters (*β* = 1 nearly unchanged), initial values, as well as the population dynamics of quarantined cases and infected cases during best fitting. From Fig. 3a-b, to produce the same outcome, the protection rate *α* and the reciprocal of the quarantine time *δ*^*−*1^ are both decreasing with the latent time *γ*^*−*1^, which is consistent with the fact that longer latent time requires longer quarantine time. Meanwhile, the initial values of exposed cases and infectious cases are increasing with the latent time. Since *E*_0_ and *I*_0_ include asymptomatic patients, they both should be larger than the number of quarantined cases. Furthermore, as the time period between the starting date of our simulation (Jan. 20th) and the initial outbreak of COVID-19 (generally believed to be earlier than Jan. 1st) is much longer than the latent time (3-6 days), *E*_0_ and *I*_0_ have to be close to each other, which makes only their sum *E*_0_ +*I*_0_ matters during the fitting. An additional important finding is that in all cases *β* is always very close to 1, which agrees with the observation that COVID-19 has an extremely strong infectious ability. Nearly every unprotected person will be infected after a direct contact with the COVID-19 patients^5,33,34^.

**FIG. 3.**
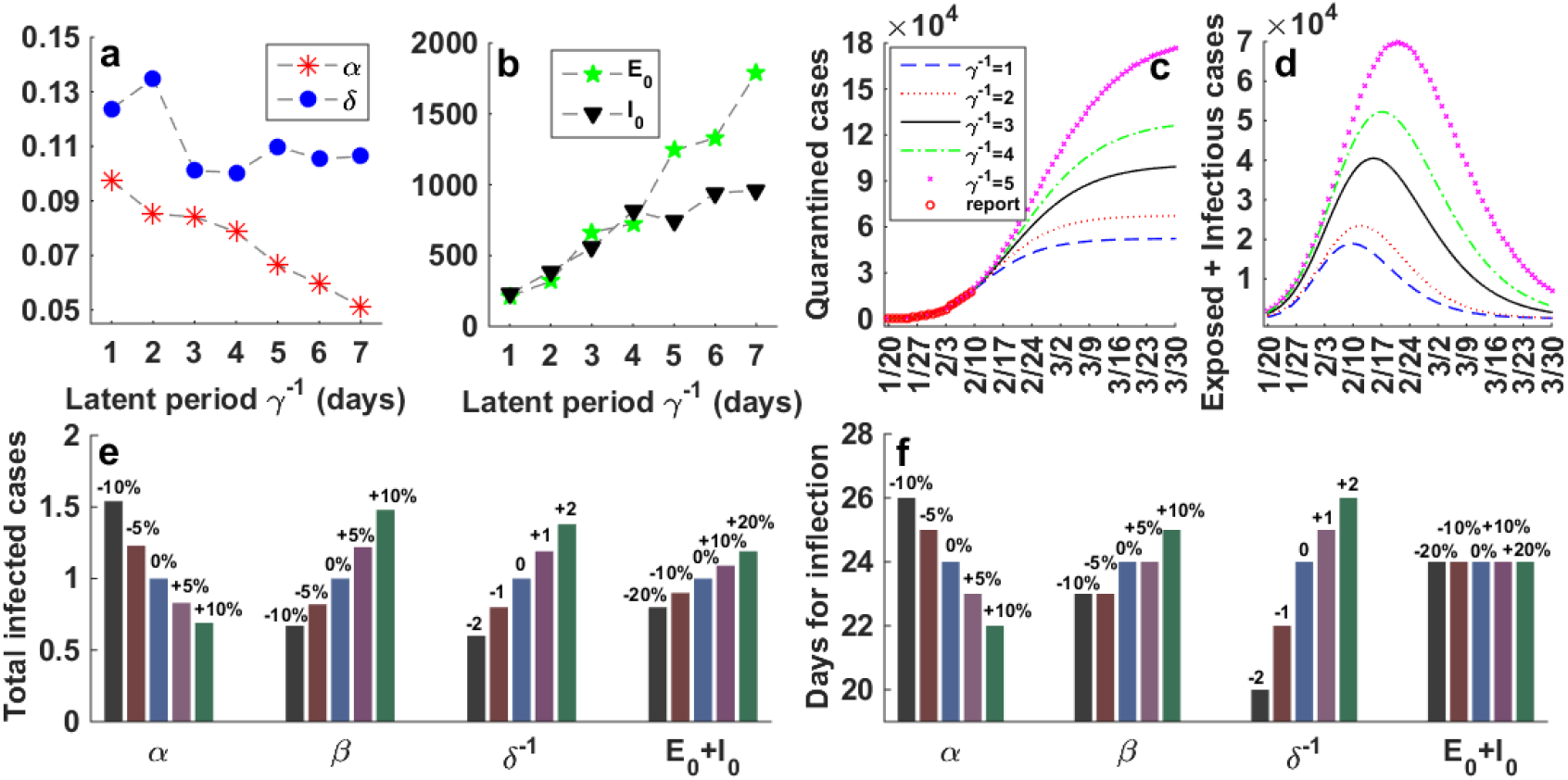
(Color online) Sensitivity analysis on parameters for the generalized SEIR model. The influence of the latent time on (a) the protection rate *α* and quarantine time *δ*^*−*1^, (b) the initial values of exposed cases *E*_0_ and infected cases *I*_0_ on Jan. 20th, (c) the cumulative quarantined cases, (d) the sum of exposed and infectious cases *E*(*t*) + *I*(*t*), *i*.*e*., the currently infected but not yet quarantined cases. (e) Effects of other parameters on the final total infected cases; (f) and the time period from the starting point (Jan. 20th) to the inflection point (when the basic reproduction number becomes less than 1). In the top panel, the value of latent time *γ*^*−*1^ is varied; while in the bottom panel, *γ*^*−*1^ is fixed as 2. All calculations are performed with respect to the data of Wuhan city, with reported data (red circles) obtained from NHC of China from Jan. 20th to Feb. 9th, 2020.

As a summary, we conclude that once the latent time *γ*^*−*1^ is fixed, the fitting accuracy on the time series data {*Q*(*t*)} basically depends on the values of *α, δ*^*−*1^ and *E*_0_ + *I*_0_. And based on a reasonable estimation on the total number of infected cases (see Fig. 3c-d), the latent time is finally determined as 2 days.

### C. Sensitivity analysis

In order to further evaluate the influence of other fitting parameters on the long-term forecast, we perform sensitivity analysis on the data of Wuhan (results for other regions are similar and not shown) by systematically varying the values of unknown coefficients^35,36^. As shown in Fig. 3e-f, the predicted total infected cases at the end of epidemic, as well as the the inflection point, at which the basic reproduction number is less than 1^6^, both show a positive correlation with the infection rate *β* and the quarantined time *δ*^*−*1^ and a negative correlation with the protection rate *α*. These facts agree with the common sense and highlight the necessity of self-protection (increase *α* and decrease *β*), timely disinfection (increase *α* and decrease *β*), early quarantine (decrease *δ*^*−*1^), *etc*. An exception is found for the initial total infected cases. Although a larger value of *E*_0_ + *I*_0_ could substantially increase the final total infected cases, it shows no impact on the inflection point, which could be learnt from the formula of basic reproduction number.

## 3. RESULTS AND DISCUSSION

### A. Interpretation of the public data

We apply our pre-described generalized SEIR model to interpret the public data on the cumulative numbers of quarantined cases, recovered cases and closed cases from Jan. 20th to Feb. 9th, which are published daily by NHC of China since Jan. 20th. Our preliminary study includes five different regions, *i*.*e*. the Mainland*, Hubei*, Wuhan, Beijing and Shanghai.

Through extensive simulations, the optimal values for unknown model parameters and initial conditions, which best explain the observed cumulative numbers of quarantined cases, recovered cases and closed cases (see Fig. 4), are determined and summarized in Table 1. There are several remarkable facts could be immediately learnt from Table 1. Firstly, the protection rate of Wuhan is significantly lower than other regions, showing many infected cases may not yet be well quarantined until Feb. 9th (the smaller *α* for Wuhan does not necessarily mean people in Wuhan pay less attention to self-protection, but more likely due to the higher mixing ratio of susceptible cases with infectious cases). Similarly, although the average protection rate for Hubei* is higher than that of Wuhan, it is still significantly lower than other regions. Secondly, the quarantine time for Beijing and Shanghai are the shortest, that for Mainland* is in between. Again, the quarantine time for Wuhan and Hubei* are the longest. Finally, the estimated number of total infected cases on Jan. 20th in five regions are all significantly larger than one, suggesting the COVID-19 has already spread out nationwide at that moment. We will come back to this point in the next part.

**TABLE I.** Summary of all constant parameters for the generalized SEIR model. E0 and I0 denote the initial values for exposed cases and infectious cases separately. The time-dependent cure rate λ(t) and mortality rate κ(t) can be read out from Fig. 2 and are given in SI.

|  | **Mainland\***<br>(exclude Hubei) | **Hubei\***<br>(exclude Wuhan) | **Wuhan** | **Beijing** | **Shanghai** |
| --- | --- | --- | --- | --- | --- |
| **Population(million)** | **1340** | **45** | **14** | **22** | **24** |
| **Protection rate ($\alpha$)** | **0.172** | **0.133** | **0.085** | **0.175** | **0.183** |
| **Infection rate ($\beta$)** | **1.0** | **1.0** | **1.0** | **0.99** | **1.0** |
| **Latent time ($\gamma^{-1}$)** | **2** | **2** | **2** | **2** | **2** |
| **Quarantine time ($\delta^{-1}$)** | **6.6** | **7.2** | **7.4** | **5.7** | **5.6** |
| **Exposed case ($E_0$)** | **696** | **592** | **318** | **27** | **18** |
| **Infectious case ($I_0$)** | **652** | **515** | **389** | **19** | **23** |

**FIG. 4.**
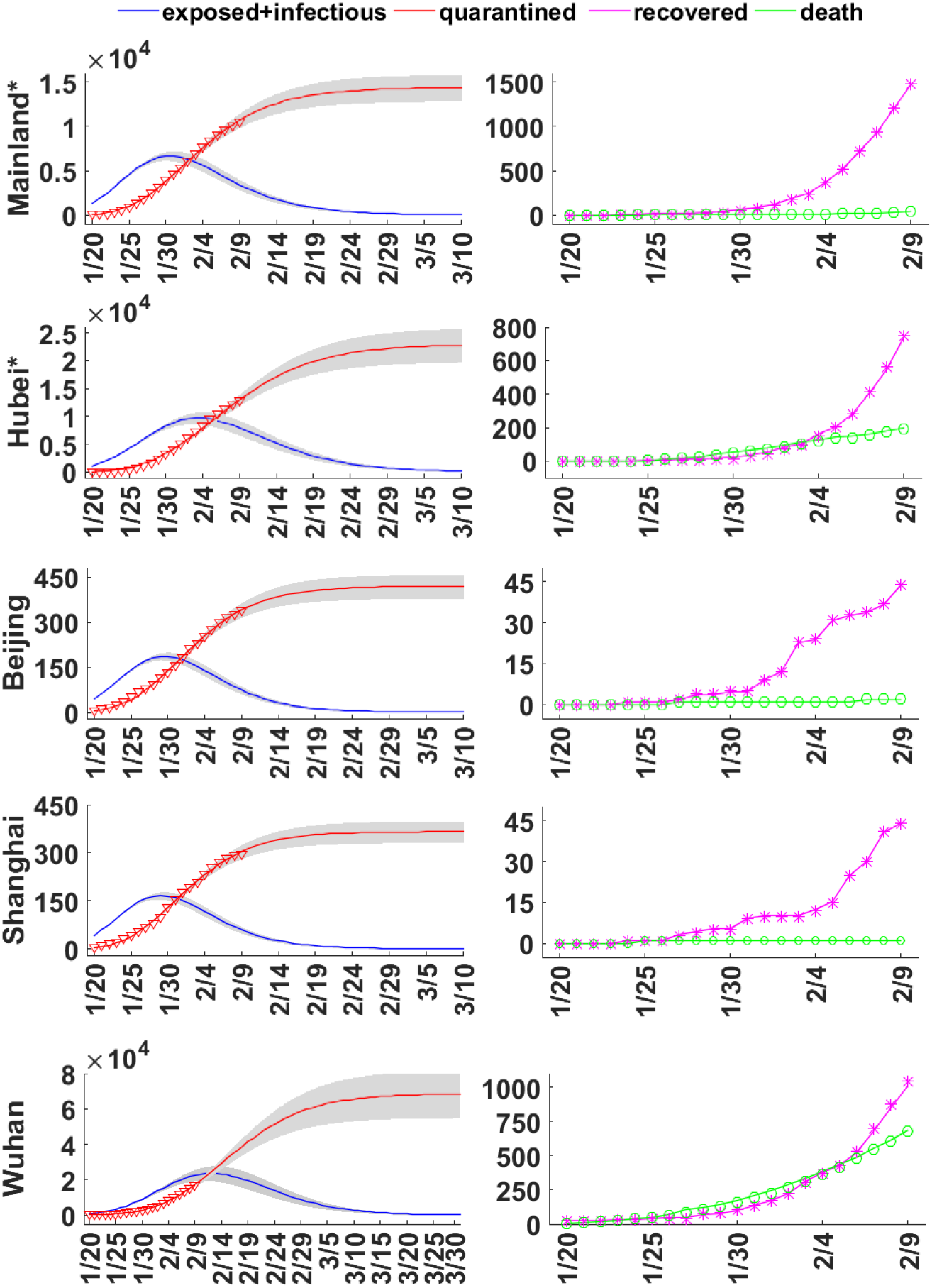
(Color online) Predictions of the generalized SEIR model on the cumulative quarantined cases (red solid lines), sum of current exposed and infectious cases (blue solid lines), cumulative recovered cases (purple solid lines), and cumulative closed cases (green solid lines) in Mainland*, Hubei*, Beijing, Shanghai, and Wuhan (from top to bottom). The red triangles, purple asterisks and green circle represent the public data points between Jan. 20th and Feb. 9th, 2020. The shaded area indicates predictions within 95% confidence interval. With the Euclidean distance ∥·∥_2_, the average relative error 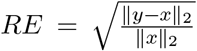 between the prediction *y* and public data *x* is evaluated for the cumulative quarantined cases, that is *RE* = 2.4%, 5.6%, 1.9%, 2.9%, 3.8% for Mainland*, Hubei*, Beijing, Shanghai and Wuhan. Parameters are taken in accordance with Table 1.

### B. Forecast for the epidemic of COVID-19

Most importantly, with the model and parameters in hand, we can carry out simulations for a longer time and forecast the potential tendency of the COVID-19 epidemic. In Fig. 4 and Fig. 5a-b, the predicted cumulative number of quarantined cases and the current number of exposed cases plus infectious cases are plotted for next 30 days as well as for a shorter period of next 13 days. Official published data by NHC of China from Feb. 10th to 15th are marked in red spots and taken as a direct validation. Overall, except Wuhan, the validation data show a well agreement with our forecast and all fall into the 95% confidence interval (shaded area). And we are delighted to see most of them are lower than our predictions, showing the nationwide anti-epidemic measures in China come into play. While for Wuhan city (and also Hubei province), due to the inclusion of suspected cases with clinical diagnosis into confirmed cases (12364 cases for Wuhan and 968 cases for Hubei* on Feb. 12th) announced by NHC of China since Feb. 12th during the preparation of our manuscript, there is a sudden jump in the quarantined cases. Although it to some extent offsets our original overestimates, it also reveals the current severe situation in Wuhan city, which requires much closer attention in the future.

**FIG. 5.**
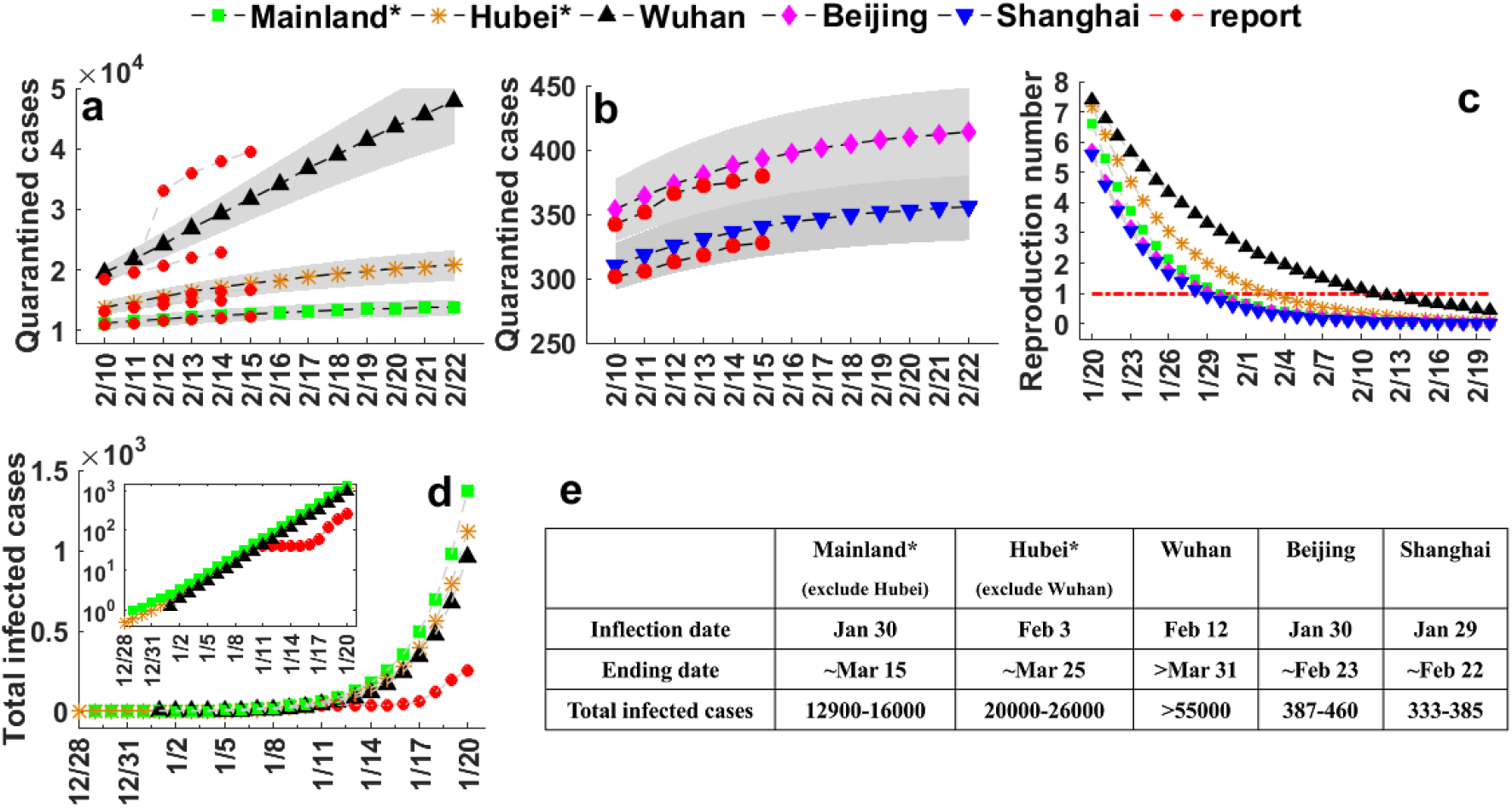
(Color online) (a-b) Predicted cumulative quarantined cases in the near future from Feb. 10th to Feb. 22nd, 2020. The shaded area indicates a 95% confidence interval. The red spots represent the reported data of Wuhan from Feb. 10th to Feb. 15th, 2020 as a validation. Parameters are taken in accordance with Table 1. (c) The basic reproduction number, (d) the estimated total infected cases at the early stage of COVID-19 epidemic between Dec. 28th, 2019 and Jan. 20th, 2020 by inverse inference, and (e) a summary on the estimated inflection point, ending date and number of final total infected cases in Mainland*, Hubei*, Wuhan, Beijing and Shanghai.

Towards the epidemic of COVID-19, our basic predictions are summarized as follows:

1. Based on optimistic estimation, the epidemic of COVID-19 in Beijing and Shanghai would soon be ended within two weeks (since Feb. 15th). While for most parts of mainland, the success of anti-epidemic will be no later than the middle of March. The situation in Wuhan is still very severe, at least based on public data until Feb. 15th. We expect it will end up at the beginning of April.
2. The estimated final total infected cases (not only total quarantined cases) for Beijing and Shanghai will be around four hundred. This number is about 13-16 thousand for mainland (exclude Hubei province), 20-26 thousand for Hubei province (exclude Wuhan city) and 55 thousand for Wuhan city.
3. According to the basic reproduction number, the inflection date for Beijing, Shanghai, mainland (exclude Hubei) is around Jan. 30th, which is close to the reported Feb. 3rd for the last one based on daily new confirmed cases. The inflection point for Hubei province (exclude Wuhan city) agrees with the reported Feb. 5th. These facts indicate that the epidemic is now under control in most cities in China.
4. The inflection point for Wuhan city is determined as Feb. 12th (data after Feb. 9th are not included into parameter estimation). By coincidence, on the same day, we witnessed a sudden jump in the number of confirmed cases due to a relaxed diagnosis caliber, meaning more suspected cases will receive better medical care and have much lower chances to spread virus. Besides, Wuhan local government announced the completion of community survey on all confirmed cases, suspected cases and close contacts in the whole city.

### C. Inverse inference on the epidemic of COVID-19

Besides the forecast, the early trajectory of the COVID-19 outbreak is also critical for our understanding on its epidemic as well as future prevention. To this end, by adopting the shooting method, we carry out inverse inference to explore the early epidemic dynamics of COVID-19 since its onset in Mainland*, Hubei*, and Wuhan (Beijing and Shanghai are not considered due to their too small numbers of infected cases on Jan. 20th). With respect to the parameters and initial conditions listed in Table 1, we make an astonishing finding that, for all three cases, the outbreaks of COVID-19 all point to 20-25 days before Jan. 20th (the starting date for public data and our modeling). It means the epidemic of COVID-19 in these regions is no later than Jan. 1st (see Fig. 5d), in agreement with reports by Li *et al*.^5,33,34^. And in this stage (from Jan. 1st to Jan. 20th), the number of total infected cases follows a nice exponential curve with the doubling time around 2 days. This in some way explains why statistics studies with either exponential functions or logistic models could work very well on early limited data points. Furthermore, we notice the number of infected cases based on inverse inference is much larger than the reported confirmed cases in Wuhan city before Jan. 20th.

## 4. CONCLUSION

In this study, we propose a generalized SEIR model to analyze the epidemic of COVID-19, which was firstly reported in Wuhan last December and then quickly spread out nationwide in China. Our model properly incorporates the intrinsic impact of hidden exposed and infectious cases on the entire procedure of epidemic, which is difficult for traditional statistics analysis. A new quarantined state, together with the recovery state, takes replace of the original *R* state in the classical SEIR model and correctly accounts for the daily reported confirmed infected cases and recovered cases.

Based on detailed analysis of the public data of NHC of China from Jan. 20th to Feb. 9th, we estimate several key parameters for COVID-19, like the latent time, the quarantine time and the basic reproduction number in a relatively reliable way, and predict the inflection point, possible ending time and final total infected cases for Hubei, Wuhan, Beijing, Shanghai, *etc*. Overall, the epidemic situations for Beijing and Shanghai are optimistic, which are expected to end up within two weeks (from Feb. 15th, 2020). Meanwhile, for most parts of mainland including the majority of cities in Hubei province, it will be no later than the middle of March. We should also point out that the situation in Wuhan city is still very severe. More effective policies and more efforts on medical care and clinical research are eagerly needed. We expect the final success of anti-epidemic will be reached at the beginning of this April.

Furthermore, by inverse inference, we find that the outbreak of this epidemic in Mainland, Hubei, and Wuhan can all be dated back to 20-25 days ago with respect to Jan. 20th, in other words the end of Dec. 2019, which is consistent with public reports. Although we lack the knowledge on the first infected case, our inverse inference may still be helpful for understanding the epidemic of COVID-19 and preventing similar virus in the future.

## Data Availability

This article used only public data.

## ACKNOWLEDGMENT

We acknowledged the financial supports from the National Natural Science Foundation of China (Grants No. 21877070, 11801020) and the Fundamental Research Funding of Beijing University of Technology (006000546318505, 006000546319509, 006000546319526). The authors would like to thank Dr. Yajing Huang for her stimulating discussions.

## AUTHOR CONTRIBUTIONS

Liu Hong designed the project. Wuyue Yang, Dongyan Zhang collected the data. All authors analyzed the data. Liangrong Peng, Wuyue Yang, Changjing Zhuge and Liu Hong wrote the manuscript, and all authors reviewed it.

## APPENDIX A. SUPPLEMENTARY MATERIALS

Supplementary materials to this article can be found online.

